# Off the rails: a pilot study of daily clinic volatility in primary care

**DOI:** 10.64898/2026.09.07.26362447

**Authors:** Nate C. Apathy, Srikar Kodali, Marylyn Presutti, Neil Siegel

## Abstract

**Background:** In primary care, rising demand and shrinking capacity produce chaotic variability during clinic days, yet this daily “schedule volatility” and its drivers are rarely measured.

**Objective:** To measure primary care physicians’ (PCPs’) assessments of daily schedule volatility and its contributing factors, and to identify potential electronic health record (EHR)-derived measures to proxy these constructs.

**Design:** Observational pilot study pairing contemporaneous end-of-day text-message (SMS) surveys with matched EHR data.

**Setting:** Five primary care clinics in the University of Maryland Medical System, August to December 2025.

**Participants:** Eight PCPs with at least 50% clinical effort.

**Measurements:** Daily schedule volatility (5-point Likert rating) and contributing factors, collected by SMS after clinic close; four EHR-derived outcome measures (same-day visit closure, cumulated time to chart closure, and the presence and volume of EHR work after 7pm) and 16 contributing-factor measures spanning tardiness, no-shows and cancellations, asynchronous (“InBasket”) volume, and schedule structure. PCP-rated volatility was dichotomized (top two “difficult-to-manage” categories vs. others) and compared using bivariate tests.

**Results:** Of 150 surveys, 105 were completed (70% response rate; median response 27 minutes). Difficult-to-manage days were uncommon (14% of days), although patient tardiness, no-shows, long-running visits, and InBasket burden were each reported on >40% of days. Difficult days involved more evening EHR work (55% vs. 19%; p=0.033), higher asynchronous volume (median 13 vs. 4 patient-message encounters; p=0.022), and higher visit volume (median 17 vs. 7; p=0.001). No-shows and same-day cancellations were not associated with volatility.

**Limitations:** Single-system pilot; bivariate, cross-sectional, and hypothesis-generating. Conclusion: PCPs’ experience of chaotic clinic days is measurable and appears driven more by asynchronous burden and visit volume than by patient no-shows or tardiness; evening EHR work is a promising scalable proxy.

**Primary Funding Source:** American Medical Association.

## Introduction

In recent years, demands on primary care providers (PCPs) in the US have shifted dramatically. In the face of flat or decreasing reimbursement rates, PCPs now bill more relative value units (RVUs) and see more visits per week on average than pre-pandemic.^1^ PCPs also provide more telemedicine, e-visits, and other forms of asynchronous (and often uncompensated) care via portal messaging.^2–5^ These demand-side increases have occurred in parallel with supply-side decreases. Historic levels of physician burnout and job dissatisfaction have led to reductions in clinical FTE and increasing rates of physician turnover, both of which put considerable pressure on the existing supply of PCPs as demand continues to rise.^6–9^ The current PCP workforce is therefore left to attempt to serve more patients in less time with fewer colleagues.

This tightening of PCP capacity amidst increasing demand manifests most clearly in PCP work schedules that often include a high volume of short, back-to-back visits coupled with double-booked appointment slots and modality-switching between in-person and telemedicine care.^10,11^ Additionally, no-shows, same-day cancellations and rescheduling, and patient tardiness are common: between 5% and 30% of ambulatory visits.^12,13^ As a result, a given clinic day can end up being unsustainably over-booked (due to double-booking, tardy patients, and/or modality-switching) or, less commonly, frustratingly under-booked (due to no-shows and same-day cancellations). This “schedule volatility” has important implications for care delivery and quality. In particular, when a physician is running behind, visits get shortened, fewer patient problems are addressed, more opioids and antibiotics are prescribed, more specialist referrals are made, and patients return more frequently for additional visits.^14–16^ Hectic and unpredictable schedules also impact physician wellbeing: when physicians feel that they do not have control over their schedules, they report lower professional fulfillment and are more likely to reduce their clinical workloads or leave practice.^8,17–19^

Given the downstream consequences of schedule volatility, both clinical and operations research has explored the predictors of patient “adherence” behavior, specifically tardiness^20–22^ and no-shows,^23–25^ including interventional studies testing strategies to reduce no-show rates.^26–29^ A separate body of literature has examined the impact of compressed clinic appointments and physician delays.^14–16,30^ Few studies, however, have linked these two streams by examining exactly how different sources of schedule disruptions translate into deteriorations or otherwise chaotic clinic days.^31^ While patient tardiness creates obvious delays to visit starts, the role of other disruptions is less clear. For example, visits also run longer than their intended time even when patients are on time, but this may be more difficult to absorb during a string of back-to-back visits than at the end of the clinic day or right before a scheduled mid-day break or reserved slot for administrative work.^32^ Similarly, double-booking may only result in delays when both patients arrive on time. Importantly, between-visit buffer time and the use of double-booking are both under the control of clinic administrators, whereas patient tardiness is not. Moreover, other clinic- or day-level factors may play a larger role: covering patients for a colleague who is out or managing lower-than-anticipated staffing both dramatically increase PCP workload and may be both more significant and more readily modifiable drivers of PCPs’ daily schedule volatility than patient behavior. However, the historical focus on patient-directed interventions has led to less understanding of how PCPs contemporaneously assess the relative impact of different schedule disruptions and their role in driving daily volatility.

The purpose of this study was to analyze PCP ratings of daily clinic schedule volatility, the core drivers of that volatility, and to explore EHR-derived measures that most closely correspond to physician perceptions of chaotic clinic days. We explore three main research questions. First, how do PCPs rate clinic days in terms of schedule volatility? Second, what factors do PCPs most commonly cite as the drivers of deviations from intended schedules? Third, what EHR-derived measures can most closely proxy both the outcome of PCP-rated volatility and PCP-reported contributing factors? Our study offers the first measurement to our knowledge of contemporaneously rated assessments of clinic day volatility and its causes. We provide both an assessment of the prevalence of highly disrupted “off the rails” clinic days and methodological innovation in the construction of measures that can reliably proxy these constructs at scale. Our findings are therefore of use to clinicians, health system and clinic administrators, and policymakers weighing different approaches to managing capacity constraints and supporting physician workforce wellbeing in primary care.

## Methods

We conducted contemporaneous PCP surveys of daily schedule volatility and analyzed matched data from the electronic health record (EHR) for 8 PCPs practicing at 5 clinics associated with the University of Maryland Medical System (UMMS) between August 2025 and December 2025. Three clinics were academic practices with residents and attending physician oversight; two were community-based, non-teaching clinics. This study was approved by the University of Maryland College Park Institutional Review Board (No. 2250527-4).

### PCP Recruitment & Questionnaire Development

We recruited PCPs via regularly held virtual departmental meetings at which a member of the research team presented the study objectives, requirements of participants, and provided links for enrollment in the study. We specifically aimed to recruit PCPs who provided clinical care for at least 50% of their work week (i.e., 50% cFTE), to ensure enough full clinic days on a weekly basis for each PCP. Attendees were also encouraged to circulate the digital study flyer and enrollment link to colleagues who were not in attendance. During the enrollment process, PCPs were asked to indicate if they would assist in survey instrument development.

The study team drafted a three-item questionnaire for review and iterative co-development with PCP study participants. We developed a brief, text-message-based set of items that could be easily completed in less than a few minutes shortly after the conclusion of a clinic day. The first item aimed to assess the PCP respondent’s overall sense of how much the day deviated from the anticipated schedule and was structured as a five-item Likert scale. PCP participants co-developed the descriptors of each response option to ensure each option was sufficiently differentiated. The second item aimed to assess the non-mutually exclusive factors that contributed the most to any deviations or volatility that occurred during that clinic day. In co-developing these response options, PCP participants determined the most common sources of schedule volatility to ensure that common options were easily selected. Participants also deemed it important to include an “Other” category for this item, with an ability to specify via free text any other factors that contributed to the daily schedule volatility that were not included in the structured response list.

### Survey Administration

During enrollment, participants provided a preferred cell phone number to facilitate text-message based surveys. Surveys were administered via Qualtrics using the SMS questionnaire functionality that allowed participants to reply to each item quickly and in relative real-time via a text message response directly in their phone’s native text message application. This approach was chosen to ensure that responses were contemporaneous with the clinic day in question. This minimizes recall bias and avoids inconveniences associated with the use of laptops, questionnaire response links, and/or survey portals requiring log-in, thus also minimizing burden to respondents.

During the survey period (August 2025 to December 2025), each PCP was sent a text-message survey on no more than one clinic day per week, also to reduce burden. To identify days the PCP was working in the clinic, the study team extracted and analyzed forward-looking schedule data from the UMMS Epic EHR. Because schedules can fluctuate over time, we conducted this forward-looking exercise on a weekly basis. On Mondays, the team received that week’s scheduling data and analyzed all PCP participant schedules to identify the days for each PCP with sufficient volume (generally at least 5 scheduled visits) to constitute a clinic day for that PCP. This allowed for inclusion of half-day clinics, which were relatively common in our PCP sample. For the identified “survey days,” the team assessed when the final visit of the day was scheduled to end, and survey text messages were scheduled to be sent within 15 minutes of the conclusion of the final visit of the day. An illustrative example of the survey interaction that includes the questionnaire language is provided in **Figure 1**.

**Figure 1.**
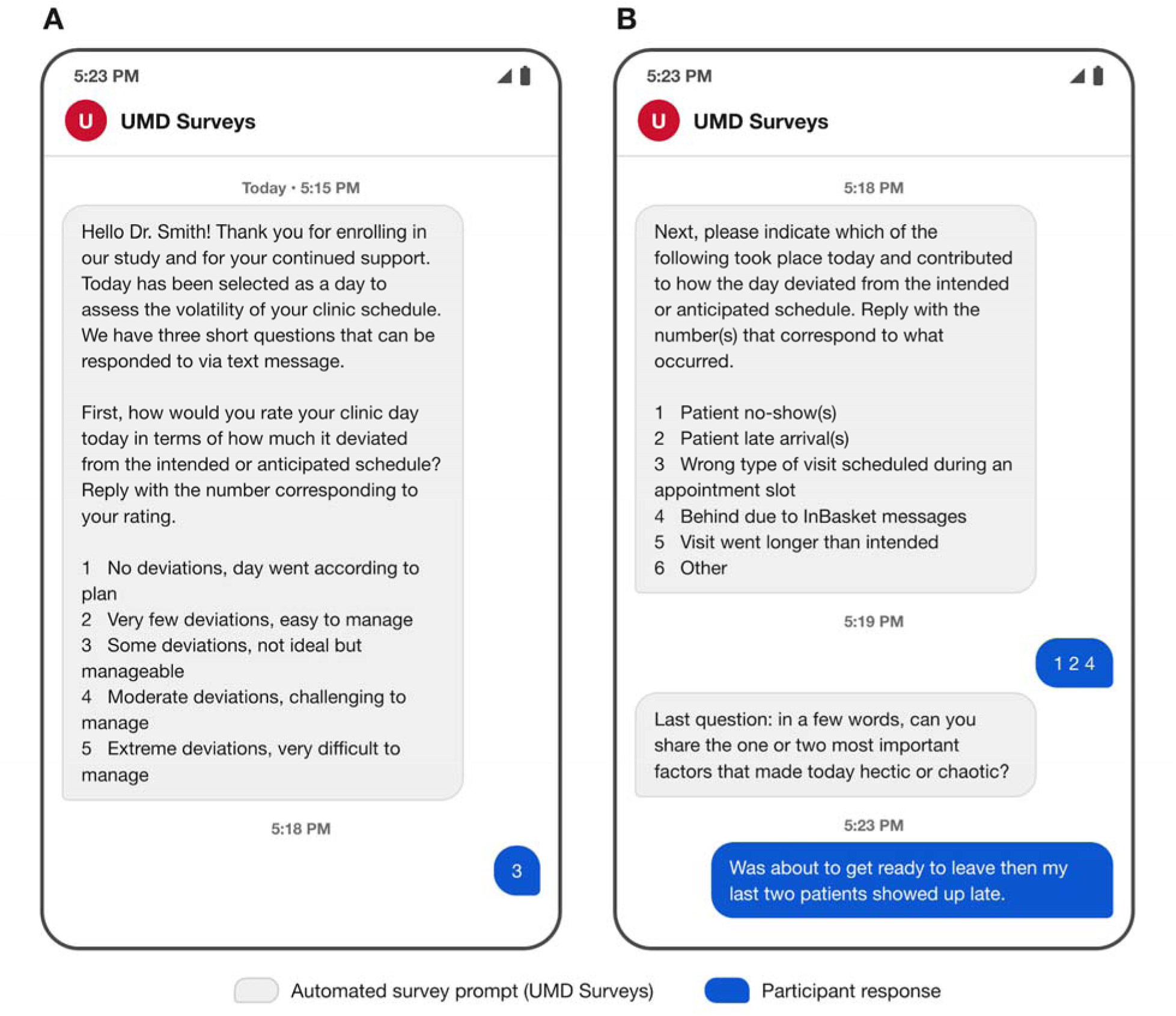
Example of text-message schedule volatility questionnaire Illustrative example of the contemporaneous, text-message-based survey sent to primary care physicians (PCPs) shortly after the end of a clinic day. (A) The first item asked the PCP to rate the day’s overall deviation from the intended schedule on a 5-point scale. (B) The second item asked the PCP to select all schedule disruptions that contributed to the day (multiple selections allowed), and the third item elicited a brief free-text description of the one or two most important contributors. Grey bubbles are automated survey prompts; blue bubbles are the participant’s replies. The physician name and message content are illustrative and do not depict a real participant. SMS = short message service.

### EHR Derived Measures: Outcomes

We used a combination of administrative and EHR audit log data to construct scalable measures of PCP-day outcomes reflecting the impacts of chaotic clinic days and clinic day characteristics. We constructed four outcome measures based on past literature and PCPs’ indications of the most consequential negative outcomes of chaotic clinic days that they shared during study enrollment. We calculated the share of completed visits that were closed on the same day as the visit;^33,34^ the median number of hours between the scheduled end of the visit and visit closure (“cumulated time to chart closure”);^35^ a binary indicator of whether or not the PCP had any evening EHR work (after 7pm) on that day;^36^ and the number of minutes of evening EHR work the PCP had, set to zero for days with no evening EHR work.^37^

### EHR Derived Measures: Contributing Factors

To characterize the factors contributing to clinic day volatility, we constructed 17 measures across four key constructs. To capture patient tardiness, we constructed four measures: a binary measure of whether the first scheduled patient of the day arrived late (i.e., check-in timestamp was after the scheduled visit start time); a binary measure of whether any patients that day arrived late; a count measure of the total number of late patients (to capture extensive margin of tardiness); and the sum of minutes by which late patients were late (to capture intensive margin of tardiness accumulation throughout the day).

To capture patient no-shows and late-breaking cancellations, we constructed four measures: a binary measure of whether the first scheduled patient of the day was a no-show; a binary measure of whether any patients that day were no-shows; a count of patients that were no-shows that day; and a count of same-day visit cancellations (i.e., cancelled date was the same as the scheduled visit date).

To capture InBasket and other asynchronous care volume, we constructed four count measures: the number of encounters denoted as “Patient Message” encounters; the number of encounters denoted as “Refill” encounters; the number of encounters denoted as “Results Follow-Up” encounters; and the number of encounters denoted as “Telephone” encounters. We excluded from these counts any encounters for patients with completed office visits on the same day. These measures do not capture the universe of message types a PCP might receive in a day, but rather those that are converted into discrete “encounters” in the EHR.

Finally, to capture structural characteristics of the schedule, we constructed five measures: total number of completed office visit and telemedicine encounters; total number of same-day add-on visits; a three-level variable capturing if the clinic day involved a) only morning visits, b) only afternoon visits, or c) both morning and afternoon visits; a binary measure of whether any time slots were double-booked; and finally a measure of “buffer time” defined as the proportion of the scheduled day not occupied by scheduled visits. We computed this measure by dividing the total number of scheduled minutes by the time span between the first scheduled visit start and last scheduled visit end time for the day and subtracting from one, such that higher proportions indicate days with greater relative buffer or break time between visits.

### Analysis

To address our first and second research questions, we analyzed survey responses to calculate the proportion of PCP-days that were rated at each level of volatility and the prevalence of each contributing factor. We also analyzed free-text responses and described the themes captured via free-text that were not previously included in the structured options (see **Figure 1**). For our third research question testing the degree to which EHR-derived measures correlated with survey response data, we first dichotomized the PCP-day volatility rating using a top-box approach, with the top two response options (“moderate deviations” and “extreme deviations”) set to one and all others set to zero. We then matched PCPs’ survey responses with the EHR-derived measures corresponding to that PCP-day. We first analyzed the relationship between our four constructed outcome measures and overall rating of schedule volatility, to examine whether these scalable outcomes were reliable proxies for PCP-rated schedule volatility. Second, we analyzed the relationship between our EHR-derived contributing factors and PCP-reported schedule disruptions, to examine the extent to which our EHR-derived measures proxied PCP-reported schedule disruptions. For this analysis, we matched EHR-derived measures to the relevant schedule disruption construct wherever possible (e.g., pairing our four measures of patient tardiness with PCP-reported patient late arrivals). Finally, to examine the generalizability of the EHR-derived measures computed for surveyed PCP-days, we analyzed all outcome and contributing factor measures for all PCP-days occurring during the study period, regardless of whether the day was a surveyed PCP-day. This analysis allowed us to evaluate whether the surveyed PCP-days differed systematically from all clinic days that occurred during the surveyed period. All analyses were constructed as bivariate analysis, and leveraged t-tests to recover p-values for normally distributed continuous measures (e.g., percentage measures); Fisher’s exact tests for categorical measures (e.g., binary indicators); and Kruskal-Wallis rank sum tests for non-normally distributed measures (e.g., skewed count measures). All EHR data was extracted from the UMMS Epic Clarity data warehouse, and all analyses were done in R v4.4.2.

## Results

We sent a total of 150 text-message surveys and received 105 complete and usable responses (70% response rate). The 45 invalid surveys include several sent erroneously on dates when the PCP was on leave or was not in clinic, reflecting changes to PCP schedules that occurred after the study team had scheduled survey distributions. The median response time from initial text message to completed response was 27 minutes, indicating that PCPs responded contemporaneously. This value reflects the elapsed time between when the survey was sent and when it was completed, not active time spent completing the survey.

### Physician-Reported Daily Schedule Volatility & Schedule Disruptions

The plurality of PCP-days (36%, n=38) were rated as having very few deviations that were easy to manage (**Figure 2A**). The next most reported day was one with some deviations that were not ideal but manageable (29%, n=30). Moderate and extreme deviations were reported for 11% (n=11) and 3% (n=3) of days, respectively. The remaining 22% (n=23) of PCP-days were rated as having no deviations from the scheduled plan. PCPs reported high rates of schedule disruptions. Patient tardiness, no-shows, long-running visits, and running behind due to InBasket messages were all reported on more than 40% of PCP-days (**Figure 2B**).

**Figure 2.**
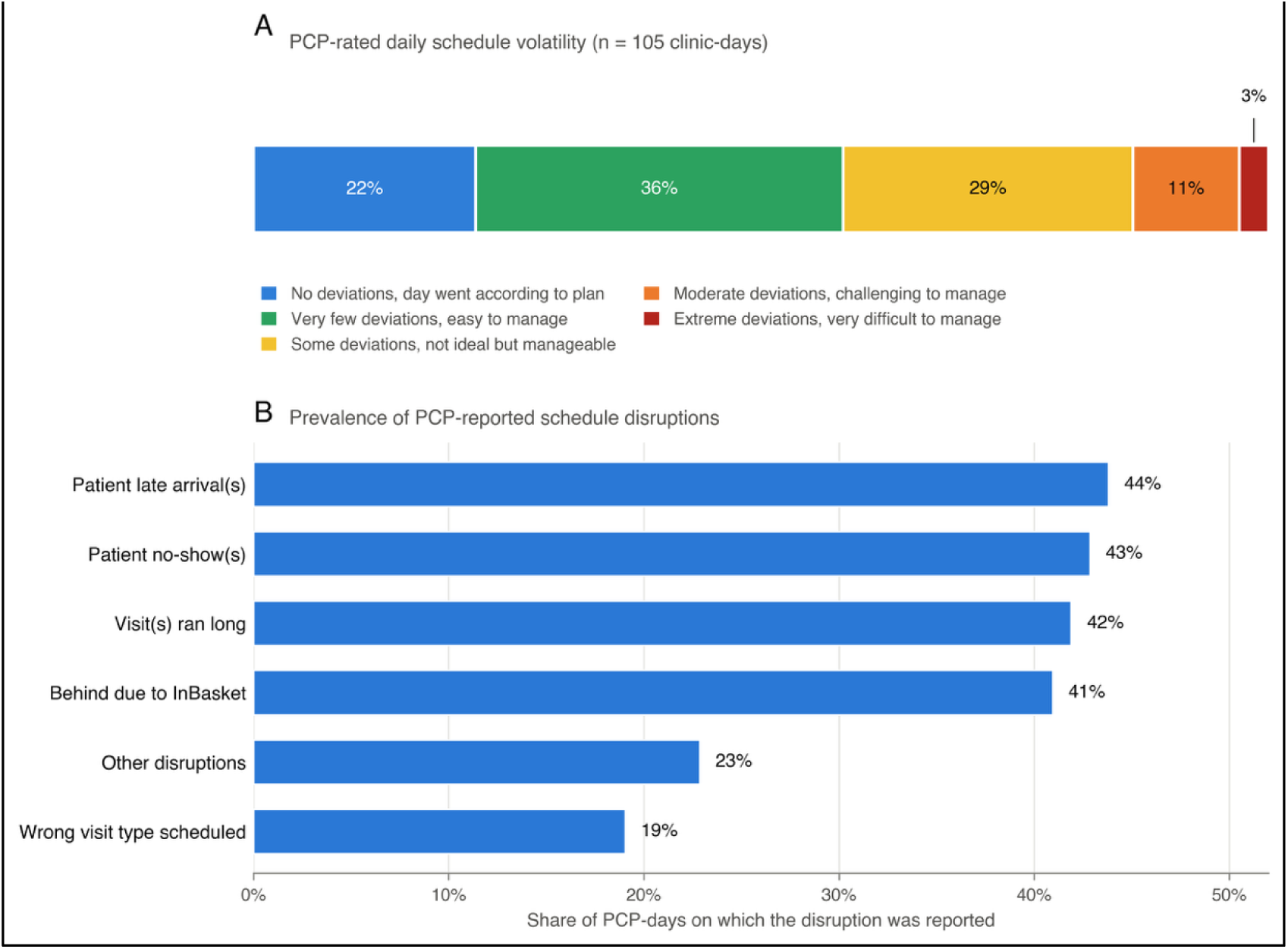
Clinic volatility ratings and schedule disruptions reported by PCPs (A) Distribution of PCP-rated daily schedule volatility across 105 surveyed clinic-days, shown as a 100% stacked bar spanning the 5-point rating scale from "no deviations, day went according to plan" (blue) to "extreme deviations, very difficult to manage" (dark red). (B) Prevalence of each PCP-reported schedule disruption, expressed as the share of the 105 clinic-days on which the disruption was reported. PCPs could report more than one disruption per day, so categories are not mutually exclusive. Percentages are rounded to whole numbers. PCP = primary care physician.

Responses to the free-text questionnaire item provided further insight into factors that contributed to the chaos of PCPs’ clinic days (**Table 1**). Common themes included *scheduling errors* (e.g., double-booking or coverage for other PCPs who were not in clinic that day); *staffing issues* (e.g., general lack of support staff or staff calling in sick); *clinical operations* (e.g., miscommunication with patients about cancellation policy, limited room availability, or slow procedures in clinic); *time management* (e.g., complex patients, unexpected clinical events, or physician practice style); and *extenuating circumstances* (e.g., after school pick-up, car troubles, family emergency, or traffic).

**Table 1.** Themes and illustrative free-text responses describing contributors to daily schedule volatility.

| **Theme** | **Illustrative free-text responses** |
| --- | --- |
| **Scheduling errors** | *"Scheduling error with one of the residents physicians, so had to accompanied more patients by double booking. Started the day with all 4 patients here waiting to be seen"*<br>*"Scheduling errors, providers absent but with full schedule"* |
| **Staffing issues** | *"Short staffed without enough medical assistants means that patients are waiting longer to get roomed and clinicians are sometimes having to perform vital signs"*<br>*"Understaffed in terms of MAs"* |
| **Clinical operations** | *"Our clinic continues to be short staffed and patients were taking longer to be roomed then is normal. Our process for administering vaccines and performing point of care tests is also slow and causes delays between patients"*<br>*"Lack of communication from front desk and not following our late policy, I'm currently waiting for my 11:20 patient that showed up 20 minutes late, automatically checked in without asking per our policy. Then it took another 20 minutes before our MA to room the patient. It's now 1205 and the patient is still not ready"* |
| **Time management** | *"Patients arriving late. Mass dumping of inbasket. Appointments running long. Need to coordinate care with other specialists between visits"*<br>*"Needed to leave on time to pick up children so pressure from that"* |
| **Extenuating circumstances** | *"Had to bring car to the shop"*<br>*"My father in law got admitted to the hospital and I had to leave clinic early"* |

Responses are verbatim free-text answers to the survey item asking PCPs to describe the one or two most important factors that made the day hectic or chaotic. PCP = primary care physician.

### EHR-Derived Outcome Measures and PCP-Reported Schedule Volatility

In the 78 survey-matched PCP clinic days with at least 3 completed visits (**Table 2**), an average of 61% of visits were closed on the same day, 24% of days involved any evening work, and the median amount of EHR work after 7pm on these days was 7.9 minutes (IQR: 1.1 to 36.1). When stratifying by PCP-rated schedule volatility, we found a counterintuitive directional relationship for efficiency measures. 72% of visits were closed same-day on difficult to manage clinic days, compared to 60% on days with manageable deviations, although this difference was not significant (p=0.36). Similarly, time to chart closure was shorter for volatile clinic days but not statistically significant (median of 0.5 hours vs. 4.0 hours, p=0.11). More intuitively, 55% of difficult to manage days involved any evening EHR work, compared to only 19% of manageable days (p=0.03). On days with evening EHR work, the median EHR time after 7pm was 26.9 minutes for difficult to manage days, compared to 7.7 minutes on manageable days (p=0.34). This large difference did not reach statistical significance given the relatively small number of days with evening EHR work.

**Table 2.** EHR-derived outcomes and contributing factors, overall and by PCP-rated daily schedule volatility.

| Measure | Full Sample (n=1,053) | Matched Survey Days (n=78) | Manageable Deviations (n=67) | Difficult to Manage (n=11) | P value* |
| --- | --- | --- | --- | --- | --- |
| **PCP-Reported Outcomes** |  |  |  |  |  |
| ***Schedule Volatility Rating*** |  |  |  |  |  |
| Manageable Deviations | — | 86% (67) | — | — |  |
| Difficult to Manage | — | 14% (11) | — | — |  |
| ***PCP-Reported Schedule Disruptions, % (n)*** |  |  |  |  |  |
| Patient late arrivals | — | 51% (40) | 48% (32) | 73% (8) | 0.226 |
| Patient no-shows | — | 50% (39) | 46% (31) | 73% (8) | 0.193 |
| Visits ran long | — | 53% (41) | 45% (30) | 100% (11) | 0.002 |
| Behind due to InBasket | — | 50% (39) | 43% (29) | 91% (10) | 0.009 |
| Wrong visit type scheduled | — | 22% (17) | 18% (12) | 46% (5) | 0.098 |
| Other disruptions | — | 19% (15) | 15% (10) | 46% (5) | 0.049 |
| **EHR-Derived Outcomes** |  |  |  |  |  |
| % of visits closed same day, mean (SD) | 81.0% (30.6) | 61% (40) | 60% (40) | 72% (41) | 0.357 |
| Hours to chart closure, median [IQR] | 2.0 [0.8, 4.9] | 3.8 [0.9, 27.2] | 4.0 [1.0, 27.9] | 0.5 [0.4, 21.7] | 0.106 |
| Any evening work, % (n) | 22.9% (241) | 24% (19) | 19% (13) | 55% (6) | 0.033 |
| Evening EHR time, min, median [IQR]† | 13.2 [0.4, 34.8] | 7.9 [1.1, 36.1] | 7.7 [0.1, 26.5] | 26.9 [10.8, 37.3] | 0.335 |
| **EHR-Derived Contributing Factors** |  |  |  |  |  |
| First patient late, % (n) | 27.2% (286) | 27% (21) | 24% (16) | 46% (5) | 0.259 |
| Any late patients, % (n) | 83.4% (878) | 95% (74) | 94% (63) | 100% (11) | 0.925 |
| No. of late patients, median [IQR] | 2.0 [1.0, 3.0] | 2.0 [1.0, 3.0] | 2.0 [1.0, 3.0] | 3.0 [2.0, 4.0] | 0.019 |
| Total late patient minutes, median [IQR] | 11.8 [3.3, 25.9] | 16.2 [7.1, 30.8] | 16.1 [7.3, 30.3] | 16.2 [6.9, 42.2] | 0.741 |
| First appointment no-show, % (n) | 11.1% (117) | 10% (8) | 12% (8) | 0% (0) | 0.501 |
| Any no-shows, % (n) | 59.4% (626) | 60% (47) | 57% (38) | 82% (9) | 0.213 |
| No. of no-shows, median [IQR] | 1.0 [0.0, 2.0] | 1.0 [0.0, 2.0] | 1.0 [1.0, 2.0] | 1.0 [1.0, 1.5] | 0.275 |
| No. of same-day cancellations, median [IQR] | 1.0 [0.0, 1.0] | 1.0 [0.0, 1.0] | 1.0 [0.0, 1.0] | 1.0 [0.5, 1.5] | 0.240 |
| No. of patient message encounters, median [IQR] | 4.0 [2.0, 8.0] | 4.0 [1.0, 7.0] | 4.0 [0.0, 7.0] | 13.0 [3.5, 13.5] | 0.022 |
| No. of medication refill encounters, median [IQR] | 6.0 [1.0, 10.0] | 5.0 [1.0, 10.0] | 4.0 [1.0, 9.0] | 10.0 [6.0, 15.5] | 0.012 |
| No. of results follow-up encounters, median [IQR] | 1.0 [0.0, 5.0] | 1.0 [0.0, 3.8] | 0.0 [0.0, 2.5] | 3.0 [0.5, 6.0] | 0.116 |
| No. of telephone encounters, median [IQR] | 1.0 [0.0, 8.0] | 1.0 [0.0, 8.8] | 1.0 [0.0, 7.5] | 12.0 [1.0, 16.5] | 0.004 |
| No. of completed office & telemedicine visits, median [IQR] | 8.0 [5.0, 13.0] | 11.0 [5.0, 15.8] | 7.0 [4.5, 14.0] | 17.0 [13.5, 19.0] | 0.001 |
| No. of same-day add-on visits, median [IQR] | 1.0 [0.0, 2.0] | 1.0 [0.0, 1.0] | 1.0 [0.0, 1.0] | 1.0 [1.0, 2.0] | 0.124 |
| Any double-bookings, % (n) | 16.6% (175) | 18% (14) | 15% (10) | 36% (4) | 0.196 |
| % buffer time, mean (SD) | 52.4% (66.3) | 42% (49) | 44% (51) | 29% (33) | 0.355 |
| **Clinic Day Type** |  |  |  |  |  |
| Full-day clinic, % (n) | 56.1% (591) | 56% (44) | 51% (34) | 91% (10) | 0.043 |
| Morning only, % (n) | 35.9% (378) | 13% (10) | 15% (10) | 0% (0) |  |
| Afternoon only, % (n) | 8.0% (84) | 31% (24) | 34% (23) | 9% (1) |  |

Data are % (n), mean (SD), or median [IQR] as indicated in each row label. Percentages are shown as whole numbers except in the Full Sample column. "Difficult to Manage" denotes the top two volatility ratings (moderate or extreme deviations); "Manageable Deviations" denotes all other rated days. Analyses were limited to surveyed clinic days with $\geq 3$ completed visits.
\* P value compares Manageable Deviations vs. Difficult to Manage (surveyed days), from t-tests for normally distributed measures; Kruskal-Wallis tests for skewed measures; and Fisher's exact tests for categorical measures.
† Conditional on non-zero evening EHR time. EHR = electronic health record; IQR = interquartile range; PCP = primary care physician; SD = standard deviation.

### EHR-Derived Contributing Factors and PCP-Reported Schedule Volatility

One EHR-derived patient tardiness measure demonstrated a difference across ratings of daily schedule volatility: difficult to manage days had a median of 3 tardy patients (IQR: 2 to 4) compared to 2 for manageable days (IQR: 1 to 3; p=0.02; **Table 2**). None of our EHR-derived measures of no-shows or same-day cancellations demonstrated a relationship with schedule volatility. Most measures of asynchronous work, however, demonstrated strong relationships with PCP-reported volatility: difficult to manage days involved a median of 13 patient message encounters, 10 medication refill encounters, and 12 telephone encounters, compared to 4, 4, and 1 of these encounter types, respectively, on manageable days (all p-values <0.05, **Table 2**).

Finally, two structural characteristics of the clinic day demonstrated bivariate relationships with PCP-reported daily volatility. First, the median number of completed office and telemedicine visits was 17 (IQR: 13.5 to 19) on difficult to manage days and 7 (4.5 to 14) on manageable days (p=0.001). Similarly, 91% of difficult to manage days were full clinic days, compared to 51% of manageable days (p=0.043). Chaotic days were also more likely to contain double-booked time slots (36% vs. 16%, p=0.20) and had a lower proportion of clinic time available as buffer time (29% of the clinic day vs. 44%, p=0.36), but neither of these differences reached statistical significance.

### EHR-Derived Contributing Factors and PCP-Reported Contributing Factors

Our EHR-derived measures in the categories of patient tardiness as well as no-shows and cancellations illustrated only suggestive agreement with PCP-reported factors contributing to schedule volatility (**Table 3**). Asynchronous care measures, however, tracked closely with PCPs reporting being “behind due to InBasket.” PCPs had a median of 5 patient message encounters (IQR: 4 to 11) on days with InBasket as a contributing factor, compared to 1 (IQR: 0 to 6.5) on days without. Similarly, PCPs had a median of 8 (IQR: 4.5 to 12) medication refill encounters on days with reported InBasket burden, compared to 1 (IQR: 0 to 6) on days without reported burden.

**Table 3.**
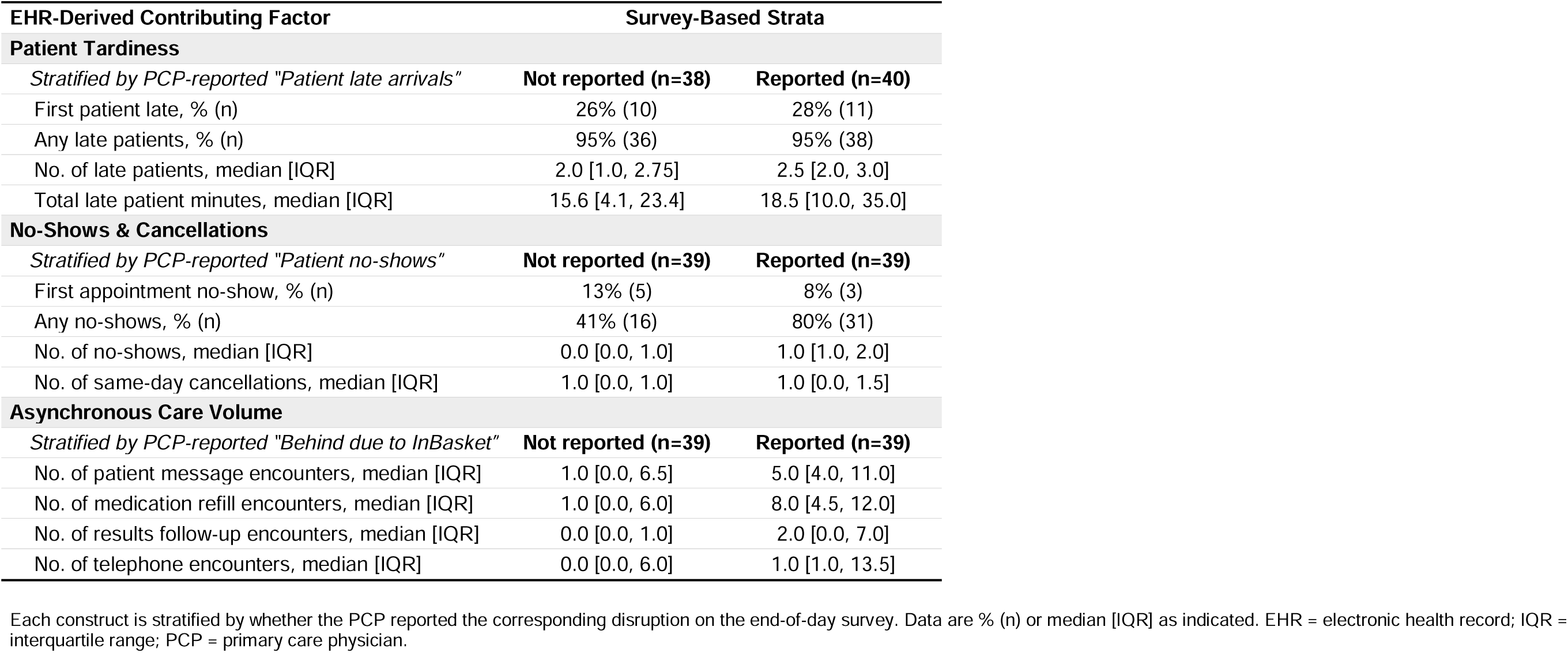
EHR-derived contributing factors stratified by corresponding PCP-reported schedule disruptions.

## Discussion

In this pilot study assessing PCP-reported schedule volatility and its concordance with EHR-derived measures, we found that highly volatile and difficult to manage clinic days were relatively rare (14% of days) and that the most significant PCP-reported contributing factors were asynchronous care burden and visits running over their allotted time. Using scalable, EHR-derived proxy outcome measures, we also found that evening EHR work (both its presence and its volume) are likely to be useful indicators of difficult-to-manage clinic days. Our findings reinforce the use of after-hours EHR time as a valuable indicator of physician well-being^36,38–41^ and extend its utility as a lagging indicator for daily volatility in ambulatory clinics. This outcome can be deployed in program evaluation to assess the success of interventions aiming to reduce schedule volatility and make ambulatory schedules less chaotic.

We also found associations between measures of asynchronous work volume and PCP-reported schedule volatility: days rated difficult to manage carried two to three times the volume of patient messages, medication refill requests, and telephone encounters as manageable days, and these measures tracked closely with PCP reports of running behind due to their InBasket. This finding is of particular import given the increase in asynchronous clinical work in the wake of the COVID pandemic.^4,42–44^ Despite the predictability, salience, and impact of this asynchronous work, dedicated time is rarely built in to PCP schedules, in part because of its “small ticket” nature and challenges associated with reimbursement for e-visits.^45–48^ But a recent evaluation found that reserving just 20 minutes in the morning and afternoon reduced after-hours EHR use by 15% with no long-term impact on revenue.^32^ Instead, most physicians must fit this work into the margins of the clinic day, where it competes directly with scheduled visits for scarce attention.^45^ Our study provides additional evidence for accounting for this asynchronous workload explicitly in physician schedules and offers support for recent proposals to create reimbursement pathways that require minimal administrative burden.^49,50^

Finally, our study constitutes a methodological contribution to the literature measuring physician work satisfaction and daily experience.^51^ To our knowledge, our study is the first to use text message surveys to capture contemporaneous assessment of PCPs’ clinic experience on a specific day. Extensions of this approach may help to reduce survey burden on clinicians (via simple, low-tech data collection), improve response rates, and improve the quality of collected data. In turn, larger scale data collection efforts can more formally validate EHR-derived measures against high-fidelity PCP self-reported data to identify the scalable, generalizable measures that can be used in program evaluation and ongoing tracking of proxy measures of physician well-being.

This single-system, small-scale pilot study has important limitations to consider. We enrolled 8 physicians and could match only 78 physician-days to complete EHR data, largely because one clinic did not permit the use of EHR data for research. This, combined with our inclusion criteria of at least three PCP-attributed completed visits, yielded a relatively small sample that was underpowered for detecting significant differences in some measures. All analyses are bivariate and cross-sectional, and do not adjust for volume or PCP characteristics (e.g., sex, age, years in practice, primary care sub-specialty, etc.), nor do we account for differences across clinics. For example, the data used for scheduling text-message surveys was unable to determine with certainty whether a given PCP-day involved oversight of trainees or consisted exclusively of visits for the PCP’s own patients, however trainees were not mentioned by any PCP respondents in any survey responses. As a result, our findings are primarily hypothesis-generating, and we make no causal claims. Two factors reported in the survey (visits running long and incorrect visit types) were not directly operationalizable from encounter data, so were not assessed with EHR-derived proxies. Finally, PCPs’ self-reported ability to manage a chaotic clinic day is subject to social-desirability bias, which may render our estimates of the prevalence of difficult-to-manage days lower than reality.

These limitations also inform future research. While discrete disruptions were commonly reported, difficult-to-manage days were relatively rare. Given this, future work should explore the role of timing of individual schedule disruptions as well as the cumulative impacts and intersecting pressures that may tip a day into chaos or prevent a day from going fully “off the rails.” This work will likely benefit from qualitative study and direct observation to both capture the many moving parts of a primary care clinic day and to conceptualize EHR-based measures that may facilitate measurement of evolving phenomena at a fine grain and at scale.

Furthermore, extensions of this work across specialties and organizations can assess the generalizability of both the survey method and the EHR-derived measures we present here. There may be specialty-specific factors that underpin schedule volatility that are distinct from those reflected in this primary care sample (e.g., patients reporting a new, unplanned issue and visits running long as a result).

Our findings have implications across the primary care enterprise. For clinicians, the asynchronous work that most sharply distinguishes chaotic days (patient messages, refills, results, and telephone encounters) is clinical labor that is largely invisible on schedules but likely warrants protected time. For clinic and health-system administrators, after-hours EHR time is a readily observed signal of which physicians and clinics may be experiencing the most volatile days. For policymakers and payers, the continued expansion of uncompensated asynchronous care strengthens the case for payment models that account for work occurring outside the visit. Rather than an unavoidable feature of modern primary care, our findings show that schedule volatility is measurable and its heaviest contributors are visible in routinely captured data.

## Data Availability

All data produced in the present study are restricted from sharing given their sensitive nature. Code to recreate similar data extracts at other sides can be made available upon reasonable request to the authors.

